# Double data extraction was insufficient for minimizing errors in evidence synthesis: a randomized controlled trial

**DOI:** 10.1101/2023.10.16.23297056

**Authors:** Lijun Tang, Ruoxi Wang, Suhail A.R. Doi, Luis Furuya-Kanamori, Lifeng Lin, Zongshi Qin, Fangbiao Tao, Chang Xu

## Abstract

**Objectives:** The objective was to investigate the role of double extraction in reducing data errors in evidence synthesis for pharmaceutical and non-pharmaceutical interventions.

**Design:** Crossover randomized controlled trial (RCT).

**Setting:** University teaching center and hospital evidence-based medicine center.

**Participants:** Eligible 100 participants were 2^nd^ year or above post-graduate students (e.g., masters, doctoral program), who were randomly (1:1) assigned for data extraction tasks of either 10 RCTs of pharmaceutical interventions or 10 of non-pharmaceutical interventions, followed by a cross-over pattern and a further double-checking process.

**Intervention:** The intervention of this trial was double-checking process for data extraction.

**Primary and secondary outcome measures:** The primary outcome was the error rates for RCTs in the pharmaceutical versus non-pharmaceutical intervention group, in terms of both study level and cell level (2 by 2 table). The secondary outcome was the absolute difference in the error rates before and after the double-checking process for both the pharmaceutical and non-pharmaceutical intervention groups, again, in terms of both study level and cell level (2 by 2 table).

**Results:** The error rates in RCTs of pharmaceutical and non-pharmaceutical groups were 64.65% and 59.90%, with an absolute difference of 4.75% and an odds ratio (OR) of 1.29 [95% confidence interval (CI): 1.06 to 1.57, *P* = 0.01] when measured at the study level. After double-checking, the error rates decreased to 44.88% and 39.54%, and the difference between the two groups remained at 5.34%, with the OR of 1.27 (95%CI: 1.1 to 1.46; *P* < 0.01). Similar results were observed when measured at the cell level.

**Conclusion:** Double-checking reduced data extraction errors, but the error rate still remained high after the process. Further evidence synthesis research may consider to use triple data extraction or else effective methods to minimize potential errors.

**Trial registration number:** Chinese Clinical Trial Registry Center (Identifier: ChiCTR2200062206)

**Strengths and limitations of the study:** This is the third randomized trial focusing on data extraction strategies and the first one in the Asia-Pacific region.

This is the first randomized trial that compares error rates of data extraction in trials of pharmaceutical interventions and non-pharmaceutical interventions.

We validated the effectiveness of double data extraction while also identifying its limitations, providing valuable evidence for future data extraction strategies.

To ensure the feasibility of the trial, we restricted subjects to 2^nd^ post-graduate students or above, which may affects the representativeness of the sample.

The readability of the chosen randomized controlled trials for pharmaceutical and non-pharmaceutical interventions may vary, potentially resulting in selection bias that can distort the outcomes.

## Introduction

Evidence synthesis is a valid method to summarize existing data of appropriate studies for a certain topic, in order to provide comprehensive, and probably, more precise evidence to support informed decision-making [1]. In evidence synthesis research, standard procedures are designed and promoted to ensure the transparency, reproductivity, and validity of the evidence bodies [2]. Among which, data extraction serves as the most important procedure which transforms the ‘raw materials’ to the new research that largely determines the reliability of the synthesized evidence [3]. However, due to the time- and labor-intensive nature, the procedure is susceptible to human errors, which eventually resulted in data extraction errors [4]. In three large-scale reproducibility studies, data extraction errors occur in almost 66.8% of meta-analyses and 85.1% of systematic reviews [5–7].

To address this issue, great efforts have been proposed to minimize potential errors during the data extraction process. A well-recognized strategy is the double data extraction, in which two reviewers extract the data independently with a consensus procedure for solving potential disagreements [8]. This method has been documented and highlighted in most of the current guidelines and collaboration groups [9–12].

There were two trials which compared the error rates of double versus single data extraction. The first trial was published in 2006 by Buscemi et al, and reported that double extraction resulted in fewer errors than single extraction with a second reviewer verification (14.5% vs. 17.7%) [13]. The second trial was published in 2018 by Li et al, and double extraction versus regular verification (single extraction plus verification) was compared. Their results suggested no substantial difference (15% vs. 16%) in the frequency of data extraction errors between these two methods [14]. These two trials both suggested minimal benefit for double data extraction and raised doubts about whether there is benefit for the extra effort expended in double extraction. Moreover, several empirical studies suggested that error rates may differ between pharmaceutical and non-pharmaceutical interventions [5, 6], and whether there is an advantage of double data extraction in bridging such differences remains unclear.

To address the above, this randomized trial attempts to provide further evidence on the role of double data extraction on reducing data extraction errors.

## Methods

### Protocol and ethical approval

This trial was approved by the Institutional Review Board at Anhui Medical University (No. 83220405), and has been registered with the Chinese Clinical Trial Registry Center (Identifier: ChiCTR2200062206). The protocol for the trial was published previously [15], and findings are reported following the CONSORT 2010 statement and its extension to randomized crossover trials [16].

### Trial design and setting

The trial was a crossover, multicenter, investigator-blinded, 1:1 randomized controlled trial, examining the data extraction error rates in RCTs of pharmaceutical and non-pharmaceutical interventions. The trial aimed to examine the role of double data extraction on reducing potential errors, and the role of double data extraction on bridging the gap in error rates between RCTs of pharmaceutical and non-pharmaceutical interventions. This trial was conducted in three centers in China, i.e, Anhui Medical University, Taihe Hospital, and Guizhou Provincial People’s Hospital. All of these centers have teaching programs about evidence-based medicine for post-graduate students. The trial was conducted in a classroom setting in every center equipped with sufficient space to accommodate up to 100 subjects, with each participant required to bring their laptop for data extraction.

This trial consisted of three stages (Figure 1). In the ***first stage***, eligible subjects were 1:1 randomly allocated into two groups (Group A and Group B) by the independent randomization center. The center sent an email to each eligible subject containing the grouping information and related materials (i.e., data extraction form and PDF files of 10 pharmaceutical or non-pharmaceutical intervention studies) by a computer-generated randomization sequence, 15 minutes before the formal trial. Subjects in Group A extracted data from 10 pharmaceutical intervention trial studies, while those in Group B extracted data from 10 non-pharmaceutical intervention studies. During the whole process, subjects were forbidden to discuss and communicate, independent third-party personnel were responsible for the order of the scene. Upon completion of data extraction, subjects emailed the completed data extraction form to an independent data manager. Due to blinding, the investigators were masked to the randomization sequence.

**Figure 1.** Trial flowchart

**Figure 2.** The error rate at cell level (mITT)

In the ***second stage***, subjects who completed the first stage in Group A crossed over to extract data from the 10 non-pharmaceutical intervention studies (i.e., studies extracted by Group B during the first stage), and vice versa for Group B.

In the ***third stage*** (double-checking period), subjects in Group A and Group B were assigned to a random number (1 to total number of subjects) in their group, and then subjects with the same number were matched as a pair for the double-checking process. Subjects who failed to be matched were out of the double-checking process. In each pair, the two subjects cross-checked the extracted data and marked the disagreements, and a further discussion on the disagreements was taken until consensus was achieved. The final data verified by the subjects were recorded on a new sheet. Between each of the three stages, subjects were granted a minimum of 30 minute break before proceeding to the next stage.

Considering that the subjects were post-graduate students with heavy teaching and research tasks, prolonged preparation times affected the compliance rate. Hence, we conduct data extraction trials every week to secure participant adherence. Each week, newly enrolled subjects were simply randomized, and the above three stages of data extraction were carried out. Subsequently, the data was sent to an independent third-party data administrator, who finally summarized the experimental results of all the subjects in all weeks.

The RCTs used in extracting data for both groups were determined based on our well-established database [6]. This database consists of 201 systematic reviews of randomized trials with 829 meta-analyses of adverse events. All the metadata within the database were checked and meta-analyses without data extraction error which had 10 or more RCTs were selected as planned. To avoid the impact of ambiguous definitions of adverse events on data extraction, we further focused on those meta-analyses which used non-composite outcomes. Finally, the systematic review by Gillies et al [17] and the systematic review by Yuan et al [18] were selected. The first systematic review investigated harms from amoxicillin, of which, the data of the meta-analysis that comparing the risk of diarrhea was used as pharmaceutical intervention group; the second systematic review investigated the differences in adverse events between internal fixation and external fixation, of which, the data of the meta-analysis that compared the risk of infection was used as non-pharmaceutical intervention group.

### Participants

Eligible subjects were post-graduate students in the 2^nd^ year of their program or above (e.g., MD or PhD program) with a background in medicine or health sciences who were studying systematic reviews, preparing to conduct ongoing systematic reviews, or already have experience conducting a systematic review. We focused on medical post-graduate students because they play a main role in data extraction in the majority of published systematic reviews. We targeted post-graduates in the 2^nd^ year of their program or above because English is not the native language of Chinese students and senior students tend to be more experienced in reading English literature.

The recruiting information was disseminated via posters in the main buildings of the three trial centers, including the teaching building and dining hall, to ensure maximum visibility. The investigator also shared e-posters on their social media (e.g., WeChat) or in community groups to further publicize the study [15]. Interested subjects were encouraged to invite their friends to participate. Upon completion of the trial, subjects received a compensation of 150 RMB (about 22 USD or 4.5 to 7.5 USD per hour), based on the expected duration of all three stages of the trial, which was estimated at 3 to 5 hours, as determined by the findings of a pilot study. All subjects provided written informed consent.

### Intervention and control

The intervention of this trial was the double-checking scheme in the third period of the trial. The first control focuses on the data extraction error rates before the double-checking stage, i.e., the first and second period, which involved single-person data extraction with subjects allowed to self-check. The second control group was the data extraction error rate of non-pharmaceutical interventions, in comparison to the RCTs of pharmaceutical interventions.

### Outcomes

The primary outcome was the error rates for RCTs in the pharmaceutical versus non-pharmaceutical intervention group. The secondary outcome was the absolute difference of the error rates before and after the double-checking process for both the pharmaceutical and non-pharmaceutical intervention groups. For a single trial, the data to be extracted contains the number of events in intervention group, the total number of subjects in intervention group, the number of events in control group, and the total number of subjects in control group, namely the 2 by 2 summarized table. Thus, there were four cells for each trial of the data. The error rates were examined at both the trial and cell levels. The former indicates the number of trials with data extraction errors on average among the 10 trials, and the later indicates the number of cells with data extraction error on average among the 10×4 = 40 cells.

We define data extraction error as any difference in data extracted from the study and the actual recorded data in the original study. It should be noted that for a RCT, two analytic rules may generally be adopted, namely, the modified intention-to-treat (mITT) and the per-protocol (PP) analysis. The data used in these two different strategies always differs; while in data extraction process, data extractors were unaware of the difference between extracted mITT information in some trials and PP in other trials, or even mixed such information in a single trial, and this may result in a difference between the extracted data and the true data. In such a case, we treated it as ***no*** data extraction error.

### Patient and Public Involvement

No patients or members of the public were involved in the design, guidance, or interpretation of this study.

### Statistical analysis

Baseline characteristics such as age, background, and experience were summarized descriptively and a chi-squared method was used to test the balance of these characteristics between groups (i.e., pharmaceutical group and non-pharmaceutical group). For the main analysis, a generalized linear mixed model (GLMM) was used to compare the error rates for data extraction of the two groups, before and after the double-checking process [19]. The GLMM is an extension to the generalized linear model (GLM) in which the linear predictor contains random effects in addition to the usual fixed effects. The main effect of the intervention was adjusted for sex, publication experience, meta-analysis experience and trial stage. The odds ratio (OR) was used as the effect estimate by setting a logit link and binomial distribution for the response variable [20].

Considering that time used for data extraction could be an important effect moderator on error rate, we further re-analyzed the data by fitting again a generalized linear mixed model while taking link function as log under the Poisson distribution [21]. This allows us to estimate the incidence risk, which measures the error rates within a time unit (5 mins). The carry-over effect was tested by using the method reported by Senn et al. [22].

The following factors that may be associated with the error rates of data extraction were included in the model: sex (female vs. male), experience of publications (with vs. without), experience on conducting meta-analysis (with vs. without), and time used for data extraction (long vs. short, stratified by the median value). Since data extraction error happens by different mechanisms, a *post hoc* analysis was done for the role of double vs single data extraction by categorizing errors into types based on the mechanisms (see details in appendix).

For the statistical inference, we pre-defined a minimal clinical important difference of the error rate to avoid the misuse of *P* value, which we set as 5% [23]. This means that when the difference of the error rates was 5% or above, we considered this a significant difference, regardless of the *P* value. The *P* value was only used to estimate the evidence against the model hypothesis at our sample size. All the analyses were run via SAS PROC GLIMMIX (SAS version 9.4, SAS Institute Inc., Cary, NC, USA).

## Results

During the period from August 25, 2022, to September 30, 2022, a total of 156 subjects underwent eligibility assessment.Among them, 56 subjects were deemed ineligible or declined to participate. 100 subjects were randomly assigned into groups (Figure 1). One participant withdrew before the start of the sutdy.. Therefore, 99 subjects (83 females, 16 males) completed the first phase of the trial (48 vs. 51). Table S1 presents the baseline characteristics of the subjects of the two groups.

After the washout period of stage 1, one participant experienced technical issues with his/her computer, resulting in data loss and inability to proceed with the second stage. The remaining 98 subjects successfully completed the second stage of the trial (47 vs. 51). Due to the simple randomization performed each week resulting in the imbalanced assignment of intervention and control groups [24], 12 subjects failed to complete the matching process (see Appendix 4). Therefore, 43 pairs (86 subjects) were matched and participated in the cross-checking stage. In the first stage, the median time used in data extraction was 1.45 hours (IQR: 1.23 to 1.60) for the group on RCTs of pharmaceutical interventions, and 1.46 hours (IQR: 1.37 to 1.64) for the group on RCTs of non-pharmaceutical interventions. In the second stage, the median time were 1.18 hours (IQR: 0.96 to 1.33) and 1.12 hours (IQR: 1.00 to 1.25). And in total, the median time for the two stages were 1.32 hours (interquartile range, IQR: 1.10 to 1.53) and 1.33 hours (IQR: 1.12 to 1.52) respectively. The median time of double-checking stage was 0.45 hours (IQR: 0.34 to 0.61) in both groups.

### Before double-checking: pharmaceutical vs. non-pharmaceutical

The crude error rates in RCTs of pharmaceutical and non-pharmaceutical intervention groups were 64.65% and 59.90% separately when measured at the study level. The absolute difference of the errors rates of pharmaceutical over non-pharmaceutical intervention groups was 4.75%, which failed to reach the minimal clinical important difference, although the evidence against the model hypothesis was strong at this sample size (OR = 1.29, 95%CI: 1.06 to 1.57, *P* = 0.01).

When measured at cell level, the crude error rates of the two groups were 36.31% and 34.14%, again, no clinically important difference was observed between the two groups (absolute difference: 2.17%), although there was strong evidence against the model hypothesis at this sample size (OR = 1.13, 95%CI: 1.02 to 1.24, *P* = 0.02).

When we assessed the time used in data extraction for each group, the error rate of each five minutes was 4.36% in pharmaceutical group and 3.82% in non-pharmaceutical group at the study level; For the cell level, the error rate of each five minutes was 2.44% in pharmaceutical group and 2.15% in non-pharmaceutical group.

### After double-checking: pharmaceutical vs. non-pharmaceutical

The crude error rates in RCTs of pharmaceutical and non-pharmaceutical intervention groups were 44.88% and 39.54% separately when measured at the study level. The absolute difference of the errors rates of pharmaceutical over non-pharmaceutical intervention groups was 5.34%, which reached the minimal clinical important difference, and the evidence against the model hypothesis was strong at this sample size (OR = 1.26, 95%CI: 1.03 to 1.53, *P* < 0.01). There was no obvious carry-over effects of the first and second stage (see details in appendix 3).

When measured at cell level, the crude error rates of the two groups were 24.07% and 20.81%, no important difference was observed between the two groups (absolute difference: 3.26%) in clinical perspective, although again there was strong evidence against the model hypothesis at this sample size (OR = 1.21, 95%CI: 1.08 to 1.36, *P* < 0.01).

When addressed the time used in data extraction for each group, the error rate of each five minutes was 4.63% in pharmaceutical group and 3.68% in non-pharmaceutical group at the cell level; For the study level, the error rate of each five minutes was 9.67% in pharmaceutical group and 7.60% in non-pharmaceutical group.

### After vs. before double-checking

The error rates in RCTs of pharmaceutical interventions before and after double-checking were 64.65% and 44.88% separately when measured at the study level, with an absolute decrease in error rates of 19.77% after the double-checking process. For RCTs of non-pharmaceutical interventions, the error rates were 59.90% and 39.54%, with an absolute decrease in error rates of 20.36%. Both of which reached the minimal clinical important difference.

At the cell level, the error rates of RCTs of pharmaceutical interventions were 36.31% and 24.07% before and after double-checking, with an absolute decrease in error rates of 12.24% after the double-checking process. For RCTs of non-pharmaceutical interventions, the error rates were 34.14% and 20.81%, with an absolute decrease in error rates of 13.33%. Again, both reached the minimal clinical important difference.

### Subgroup and post hoc analysis

Table 1 presents the results of subgroup analysis. Specifically, data extraction errors were smaller in those with experience than those without (53.0% vs. 68.0%, OR = 0.46, 95%CI: 0.31 to 0.68, *P* < 0.01) when measured at the study level. Similar results were observed at the cell level (28.9% vs. 39.1%, OR = 0.63, 95%CI: 0.48 to 0.83, *P* < 0.01).

**Table 1.** Results of generalized linear mixed model before double-checking (mITT)

| Comparisons | OR (95% CI) | P-value |
| --- | --- | --- |
| **Intervention\*** |  |  |
| Pharmacological vs. Non-pharmacological | 1.13(95% CI: 1.02 to 1.24) | 0.02 |
| **Expertise\*** |  |  |
| 1 to 2 years vs. No | 0.82(95% CI: 0.61 to 1.07) | 0.20 |
| **Experience\*** |  |  |
| with vs. without | 0.63(95% CI: 0.48 to 0.83) | <0.01 |
| **Sex\*** |  |  |
| Female vs. Male | 0.98(95% CI: 0.75 to 1.29) | 0.89 |
| **Intervention □** |  |  |
| Pharmacological vs. Non-pharmacological | 1.29(95% CI: 1.06 to 1.57) | 0.01 |
| **Expertise □** |  |  |
| 1 to 2 years vs. No | 0.74(95% CI: 0.50 to 1.08) | 0.12 |
| **Experience □** |  |  |
| with vs. without | 0.46(95% CI: 0.31 to 0.68) | <0.01 |
| **Sex □** |  |  |
| Female vs. Male | 0.92(95% CI: 0.64 to 1.32) | 0.65 |

\*: cell level;
□: study level

**Table 2.** Results of generalized linear mixed model after double-checking (mITT)

| Comparisons | OR (95% CI) | P-value |
| --- | --- | --- |
| **Intervention\*** |  |  |
| Pharmacological vs. Non-pharmacological | 1.21(95% CI: 1.08 to 1.36) | <0.01 |
| **Expertise\*** |  |  |
| 1 to 2 years vs. No | 1.14(95% CI: 0.9 to 1.43) | 0.58 |
| **Experience\*** |  |  |
| with vs. without | 0.68(95% CI: 0.54 to 0.85) | 0.01 |
| **sex\*** |  |  |
| Female vs. Male | 1.01(95% CI: 0.81 to 1.25) | 0.73 |
| **Double-checking\*** |  |  |
| After vs. before | 0.53(95% CI: 0.44 to 0.64) | <0.01 |
| **Intervention □** |  |  |
| Pharmacological vs. Non-pharmacological | 1.26(95% CI: 1.03 to 1.53) | <0.01 |
| **Expertise□** |  |  |
| 1 to 2 years vs. No | 1.12(95% CI: 0.84 to 1.48) | <0.01 |
| **Experience□** |  |  |
| with vs. without | 0.64(95% CI: 0.49 to 0.85) | <0.01 |
| **Sex□** |  |  |
| Female vs. Male | 1.05(95% CI: 0.81 to 1.36) | 0.72 |
| **Double-checking□** |  |  |
| After vs. before | 0.40(95% CI: 0.31 to 0.51) | <0.01 |

\*: cell level;
□: study level

The results of the *post hoc* analysis were presented in Figure 3. Specifically, data extraction errors were significant higher in the numerical error than mismatching error (52.12% vs. 10.15%). After double-checking, there was a decrease in numerical error than before checking (29.54% vs. 52.12%), while a slight while unsignificant increase in mismatching error (12.67% vs. 10.15%). Similar results were observed for RCTs of pharmaceutical and non-pharmaceutical interventions.

**Figure 3.** The error rate at study level (mITT)

## Discussion

In this trial, we examined the differences in data reproducibility issues between pharmaceutical and non-pharmaceutical intervention RCTs and assessed the role of performing double--checking on error rates. Our findings suggested that extracting data from RCTs of pharmaceutical interventions, reviewers are more likely to make errors in evidence synthesis practice than from RCTs of non-pharmaceutical interventions, while the amount of increase in errors were generally not clinically significant. The outcomes of this study diverge from our prior study, which reported a twofold higher data extraction error rate in pharmaceutical interventions compared to non-pharmaceutical interventions. This variance might be attributed to the outcomes of this study focus on non-composite outcomes, in contrast to previous studies encompassing all outcomes.

In addition, double data extraction can effectively reduce data extraction error rates, with 20% absolute decrease of the error rate at the study level and 12% at the cell level. Despite this commendable decrease, it is noteworthy that the error rates still remained concerning even after the double-checking process (more than 40% and 20% at study and cell level). The remaining errors may hold the potential to impact the estimation of the estimate, and in some cases even alter the overall direction of the findings. Further evidence synthesis researches may consider to use triple data extraction to minimize potential errors.

According to our classification of data errors, the rate of numerical errors was higher than mismatching errors, aligning with the findings of our previous studies [6]. Interestingly, after undergoing double-checking process the overall error rate and numerical error rate decreased, while the mismatching error rate showed a slight increase. This may suggest that the role of double-checking process may differ for error mechanisms and it makes no sense on reducing the occurrence of mismatching error. Maybe data extractors focused more on the correctness of the numerical values while ignored whether the data matched correctly to the groups. Further systematic review authors should pay attention to such types of error during the data extraction process.

To the best of our knowledge, this is the largest RCT that investigated the role of double-checking on data extraction errors involving pharmaceutical and non-pharmaceutical interventions. Our sample size ensures the representativeness of results. When defining the data extraction error, we considered both ITT and PP rules of the RCTs, which expected to more reasonable to reflect the error rate. In addition, the template RCTs we used for data extraction tasks have been quadra-checked to ensure the accuracy, which provided a strong safeguard against potential human errors. Moreover, current study is the first to examine the role of double-checking process on error rates in terms of the type of the errors due to different mechanisms.

There might be two major limitations. First, the readability of the content of the selected RCTs on pharmaceutical interventions and non-pharmaceutical interventions may differ. This may lead to selection bias that distorted the results. While this bias could in turn dismissed by the prolonged time. In our study, we did not detect a difference of the time in the two groups and thus we expect the impact of this bias would be minor. Second, the participants of current study were all medical students, the representativeness would be limited since a proportion of systematic reviews were employed by clinicians and principal researchers. Further randomized trials focus on the clinicians or researchers for data extraction errors is worthy.

## Conclusion

When extracting data from RCTs of pharmaceutical interventions, reviewers are more likely to make errors in evidence synthesis practice than from RCTs of non-pharmaceutical interventions, while the amount of increase in errors were generally not clinically significant. Double data extraction can effectively reduce data extraction errors, however, the error rates after the double-checking process still remains a high level. Further evidence synthesis researches may consider to use triple data extraction to minimize potential errors.

## Data Availability

The data will be shared to the public after the publication of the trial within 3 months.

## Acknowledge

We gratefully acknowledge the assistance provided by Taihe Hospital; Department of pharmacological, Guizhou Provincial People’s Hospital; many other individuals on the project. We also thanks to Yi Zhu, Xi Yang, Shiqi Fan, Yuxin Zhang, Chenfang Wang, Mengya Zhao, and Xiangxin Zhang from Anhui Medical University for the help of trial management.

## Ethics

This RCT was registered on the Chinese Clinical Trial Registry (Identifier: ChiCTR2200062206). Ethics approval was obtained from the Anhui Medical University IRB (No.83220405).

## Contributors

Conception and design: CX; Manuscript drafting: LJ.T, RX.W, CX; Data collection: LJ.T, RX.W; Data analysis and result interpretation: LL, LJ.T, CX; Statistical guidance: LFK, SD; Methodology guidance: LFK, SD; Manuscript editing: YZ, XY, SD, LFK, LL, ZSQ, FBT and CX. All authors have read and approved the manuscript. All authors approved the final version to be published.

## Data sharing

The data will be shared to the public after the publication of the trial within 3 months.

## Conflict of interests

The original first author and second author has quitted from the project and no longer be listed as authors.

## Funding

This work was supported by the National Natural Science Foundation of China (72204003), the Academic Discipline Development (0301001882) from Anhui Medical University and the program grant #NPRP-BSRA01-0406-210030 from the Qatar National Research Fund (a member of Qatar Foundation). The funding bodies had no role in any process of the study (i.e., study design, analysis, interpretation of data, writing of the report, and decision to submit the article for publication).

**Funding:** This work was supported by the National Natural Science Foundation of China (72204003), the Academic Discipline Development (0301001882) from Anhui Medical University and the program grant #NPRP-BSRA01-0406-210030 from the Qatar National Research Fund (a member of Qatar Foundation).

## Notes

### Competing Interest Statement

The original first author and second author has quitted from the project and no longer listed as authors.

### Clinical Trial

ChiCTR2200062206

### Clinical Protocols

https://linkinghub.elsevier.com/retrieve/pii/S2451-8654(23)00135-7

### Author Declarations

This trial was approved by the Institutional Review Board at Anhui Medical University (No. 83220405), and has been registered with the Chinese Clinical Trial Registry Center (Identifier: ChiCTR2200062206).

## Reference

1. Djulbegovic B, Guyatt GH. Progress in evidence-based medicine: a quarter century on. Lancet 2017; 390(10092): 415–423.

2. Committee on Standards for Systematic Reviews of Comparative Effectiveness Research, Board on Health Care Services. In: Eden J, Levit L, Berg A, Morton S, editors. Finding what works in health care: standards for systematic reviews. Washington, DC: National Academies Press; 2011.

3. Mathes T, Klaßen P, Pieper D. Frequency of data extraction errors and methods to increase data extraction quality: a methodological review. BMC Med Res Methodol 2017; 17:152.

4. Pradhan R, Hoaglin DC, Cornell M, et al. Automatic extraction of quantitative data from ClinicalTrials.gov to conduct meta-analyses. J Clin Epidemiol 2019 ;105:92–100.

5. Xu C, Doi SA, Zhou XQ, et al. Data reproducibility issues and their potential impact on conclusions from evidence syntheses of randomized controlled trials in sleep medicine. Sleep Med Rev 2022 ;66:101708.

6. Xu C, Yu T, Furuya-Kanamori L, et al. Validity of data extraction in evidence synthesis practice of adverse events: reproducibility study. BMJ 2022;377: e069155.

7. Jones AP, Remmington T, Williamson PR, et al. High prevalence but low impact of data extraction and reporting errors were found in Cochrane systematic reviews. J Clin Epidemiol 2005; 58:741–742.

8. Egger M, Davey Smith G, Altman DG, editors. Systematic reviews in health care: meta-analysis in context. 2nd ed. London: BMJ Publishing Group; 2001.

9. Chandler J, Cumpston M, Thomas J, et al. Chapter I: Introduction. In: Higgins JPT, Thomas J, Chandler J, et al (editors). Cochrane Handbook for Systematic Reviews of Interventions version 6.3. Cochrane, 2022. Available from www.training.cochrane.org/handbook.

10. Shea BJ, Reeves BC, Wells G, et al. AMSTAR 2: a critical appraisal tool for systematic reviews that include randomised or non-randomised studies of healthcare interventions, or both. BMJ 2017; 358: j4008.

11. Tufanaru C, Munn Z, Aromataris E, et al. Chapter 3: Systematic reviews of effectiveness. In: Aromataris E, Munn Z (Editors). JBI Manual for Evidence Synthesis. JBI, 2020. Available from https://synthesismanual.jbi.global.

12. Page MJ, McKenzie JE, Bossuyt PM, et al. The PRISMA 2020 statement: an updated guideline for reporting systematic reviews. BMJ 2021; 372: n71.

13. Buscemi N, Hartling L, Vandermeer B, et al. Single data extraction generated more errors than double data extraction in systematic reviews. J Clin Epidemiol 2006; 59(7): 697–703.

14. Li T, Saldanha IJ, Jap J, et al. A randomized trial provided new evidence on the accuracy and efficiency of traditional vs. electronically annotated abstraction approaches in systematic reviews. J Clin Epidemiol 2019; 115: 77–89.

15. Zhu Y, Ren P, Doi SAR, et al. Data extraction error in pharmaceutical versus non-pharmaceutical interventions for evidence synthesis: Study protocol for a crossover trial. Contemp Clin Trials Commun 2023; 35: 101189.

16. Dwan K, Li T, Altman DG, et al. CONSORT 2010 statement: extension to randomised crossover trials. BMJ 2019; 366: l4378

17. Gillies M, Ranakusuma A, Hoffmann T, et al. Common harms from amoxicillin: a systematic review and meta-analysis of randomized placebo-controlled trials for any indication. CMAJ 2015; 187(1): E21–E31.

18. Yuan ZZ, Yang Z, Liu Q, et al. Complications following open reduction and internal fixation versus external fixation in treating unstable distal radius fractures: Grading the evidence through a meta-analysis. Orthop Traumatol Surg Res 2018; 104(1): 95–103.

19. Xu C, Furuya-Kanamori L, Lin L. Synthesis of evidence from zero-events studies: A comparison of one-stage framework methods. Res Synth Methods 2022; 13(2): 176–189.

20. Doi SA, Furuya-Kanamori L, Xu C, Lin L, Chivese T, Thalib L. Controversy and Debate: Questionable utility of the relative risk in clinical research: Paper 1: A call for change to practice. J Clin Epidemiol 2022;142:271–279.

21. Bagos PG, Nikolopoulos GK. Mixed-effects Poisson regression models for meta-analysis of follow up studies with constant or varying durations. Int J Biostat 2009; 5:21: article 21

22. Senn SS. Cross-over trials in clinical research. John Wiley & Sons, 2002.

23. Chan LS. Minimal clinically important difference (MCID)--adding meaning to statistical inference. Am J Public Health 2013 ;103(11): e24–5.

24. Randelli P, Arrigoni P, Lubowitz JH, et al. Randomization procedures in orthopaedic trials. Arthroscopy. 2008 ;24(7):834–8.

